# Mitigating the psychological impacts of COVID-19 restrictions on older people: The UK Behavioural Activation in Social Isolation (BASIL+) COVID-19 Urgent Public Health (UPH) trial and living systematic review

**DOI:** 10.1101/2023.06.13.23291329

**Authors:** Simon Gilbody, Elizabeth Littlewood, Dean McMillan, Lucy Atha, Della Bailey, Kalpita Baird, Samantha Brady, Lauren Burke, Carolyn A. Chew-Graham, Peter Coventry, Suzanne Crosland, Caroline Fairhurst, Andrew Henry, Kelly Hollingsworth, Elizabeth Newbronner, Eloise Ryde, Leanne Shearsmith, Han-I Wang, Judith Webster, Rebecca Woodhouse, Andrew Clegg, Sarah Dexter-Smith, Tom Gentry, Catherine Hewitt, Andrew Hill, Karina Lovell, Claire Sloan, Gemma Traviss-Turner, Steven Pratt, David Ekers

## Abstract

**Background:** Older adults were more likely to be socially isolated during the COVID-19 pandemic, with risk of depression and loneliness. Behavioural Activation (BA) could feasibly maintain mental health in the face of COVID isolation.

**Methods:** We undertook a multicentre randomised controlled trial [BASIL+ ISRCTN63034289] of BA to mitigate depression and loneliness among older people. BA was offered by telephone to intervention participants (n=218). Control participants received usual care, with existing COVID wellbeing resources (n=217).

**Findings:** Participants engaged with 5.2 (SD 2.9) of 8 remote BA sessions. Adjusted mean difference (AMD) for depression (PHQ-9) at 3 months [primary outcome] was -1.65 (95% CI -2.54 to -0.75, p<0.001). There was an effect for BA on emotional loneliness at 3 months (AMD -0.37, 95% CI -0.68 to -0.06, p=0.02), but not social loneliness (AMD -0.05, 95% CI -0.33 to 0.23, p=0.72). Other secondary outcomes at 3 months were anxiety (GAD-7: AMD -0.67, 95% CI -1.43 to 0.09, p=0.08) and quality of life (SF12 mental component: AMD 1.99, 95% CI 0.22 to 3.76, p=0.03; physical component: AMD - 0.50, 95% CI -2.14 to 1.10, p=0.53).

BASIL+ trial results were incorporated into a living systematic review [PROSPERO CRD42021298788], and we found strong evidence of an impact of behavioural and/or cognitive strategies on depression [random effects pooled standardised mean difference -0.32, 95% CI -0.48 to -0.16, 10 studies, n=1,210 participants] and loneliness [random effects pooled standardised mean difference -0.44, 95%CI -0.64 to -0.24, 13 studies, n=1,421 participants] in the short-term (<6 months).

**Interpretation:** BA is an effective intervention that reduces depression and some aspects of loneliness in the short term. This adds to the range of strategies to improve population mental health, particularly among older adults with multiple long-term conditions. These results will be helpful to policy makers in preventing depression and loneliness beyond the pandemic.

**Funding:** NIHR RP-PG-0217-20006

## Introduction

The mental health of the population deteriorated during the COVID-19 pandemic.^1^ People reported social isolation, and the incidence of depression and anxiety increased for older people and those with medical vulnerabilities.^2^ A plausible mechanism for this deterioration was that COVID-19 restrictions led to disruption of daily routines, loss of social contact and heightened isolation and increased loneliness; each of which are linked to mental ill health.

Social isolation, social disconnectedness, perceived isolation and loneliness are known to be linked to common mental health problems, such as depression in older people.^3^ Loneliness is a risk factor for depression and seems detrimental to physical health and life expectancy.^4^ It is recognised that strategies that maintain social connectedness could be important in ensuring the mental health of older people,^5^ particularly during the pandemic and in the planning for post-pandemic recovery.^6^

The need for research to mitigate the psychological impacts of COVID-19, particularly loneliness, was highlighted as a priority at the outset of the pandemic,^7^ and we responded by designing and delivering one of only a small number of multi-centre trial programmes to evaluate psychosocial interventions to maintain the mental health of vulnerable populations.^8^

Behavioural activation (BA) is an evidence-based psychological treatment that explores how physical inactivity and low mood are linked and result in a reduction of valued activity.^9^ Within BA, the therapist and patient work together to develop a collaborative treatment plan that seeks to reinstate (or replace, if former activities are no longer possible) behaviours that connect people to sources of positive reinforcement (meaningful activity), including social connectedness.

Small scale trials of BA delivered to socially-isolated older people have produced encouraging preliminary results,^10^ but at the outset of the pandemic there was not sufficient research evidence to support whole-scale adoption, or to inform the population response to COVID-19 or in planning for post-pandemic recovery.^7^ In early 2020, we therefore adapted an ongoing programme of work into the role of BA in multiple long-term conditions (‘multimorbidity’) to answer the following overarching question: **‘Can we prevent or ameliorate depression and loneliness in older people during COVID isolation?’**.

In this paper we present data up to the primary endpoint (3 months) of the BASIL+ trial (<u>B</u>ehavioural <u>A</u>ctivation in <u>S</u>ocial Isolation): a fully-powered multi-centre randomised controlled trial (RCT) of manualised BA, adapted specifically to be delivered at scale and remotely (via the telephone) for older adults who were at risk of socially isolation as a consequence of COVID-19 (including the post-pandemic period).

The results of an external pilot trial of the BASIL+ intervention have already been published ^11^ and showed a signal of effect in reducing loneliness at 3 months in the BA group relative to control [De Jong Gierveld Loneliness scale: adjusted mean difference -0.87; 95% CI -1.56 to -0.18). Evidence from a living systematic review [PROSPERO CRD42021298788]^11^ demonstrates a growing evidence base of the clinical effectiveness of cognitive and/or behavioural approaches for loneliness (standardised mean difference [SMD] -0.48, 95% CI -0.70 to -0.27) and depression (SMD -0.31, 95% CI -0.51 to -0.11). To our knowledge the BASIL+ trial is the first large-scale fully powered trial of a brief psychological intervention to mitigate loneliness, alongside other common mental health problems such as depression and anxiety.

## Methods

### Study design and participants

BASIL+ was a two arm, parallel group RCT. Here we present the primary [depression severity] and secondary outcomes up to the primary endpoint [three months post-randomisation]. The design was informed by an external developmental phase. BASIL+ study recruitment and follow up procedures were first tested in an external pilot RCT (BASIL-C19 pilot RCT)^12, 13^ with a concurrent qualitative study.^14^ The BASIL-C19 pilot trial showed that it was possible to deliver a telephone-based intervention during the COVID-19 pandemic and to achieve high levels of follow up [see ^11^ and ^12^ for a detailed report of the BASIL developmental phase].

The COVID-19 responsive BASIL trials programme was supported by the National Institute for Health and Care Research (NIHR) under grant RP-PG-0217-20006, and was adopted by the NIHR Urgent Public Health programme on 28/05/2020 (https://www.nihr.ac.uk/covid-studies/study-detail.htm?entryId=249030). The protocol and design for the BASIL+ study was registered (ISRCTN63034289) on 08/02/2021 and is available in the public domain^15^. The trial protocol is attached [supplementary file s1]. Recruitment activity took place between 08/02/2021 and 28/02/2022 (54 weeks in total).

Inclusion criteria: Based on the United Kingdom (UK) Academy of Medical Sciences definition of multimorbidity^16^ we recruited older adults (65 years or over) with two or more physical long-term conditions (LTCs) on primary care registers in twenty six general practices in England and Wales. Participants had to have a score of ≥5 on the Patient Health Questionnaire (PHQ-9), putting them at risk of clinical depression or indicating already established depressive symptoms. Participants included those that were subject to UK Government guidelines regarding COVID-19 self-isolation, social distancing and shielding as relevant to their health conditions and age (though this was not a requirement and these requirements changed several times during the study period).

Exclusion criteria: Older adults who had cognitive impairment, bipolar disorder/psychosis/psychotic symptoms, alcohol or drug dependence, were in the palliative phase of illness, had active suicidal ideation, were receiving psychological therapy, or unable to speak or understand English. Older adults were not excluded on the basis of living in residential/care homes.

Potentially eligible patients were identified and contacted by telephone by staff working with the general practices. Interested patients could also complete an online consent form or contact the study team directly.

### Randomisation, concealment of allocation and blinded assessment of outcome

After consent, eligible participants completed a baseline questionnaire over the telephone with a study researcher. Participants were then randomised and informed of their group allocation (BASIL+ intervention or usual care with signposting). Participants were allocated in a 1:1 ratio using simple randomisation without stratification. The allocation schedule was generated in Stata v16 (StataCorp. 2019. Stata Statistical Software: Release 16. College Station, TX: StataCorp LLC.) by the trial statistician, who was not otherwise involved in the recruitment of participants. Treatment allocation was concealed from study researchers at the point of recruitment using an automated computer data entry system, administered remotely by York Trials Unit, University of York. Owing to the nature of the intervention, none of the participants, general practices, study clinicians, or BASIL+ Support Workers (BSWs) could be blinded to treatment allocation. Researchers facilitating the telephone-based outcome assessments at follow up were blind to treatment allocation (‘blinded outcome assessment’).

**Intervention (Behavioural Activation):** The intervention (BA within a collaborative care framework) was adapted for the purposes of the BASIL+ trial [see^11, 12^ for a description of the COVID-19 social isolation adaptation]. BA is a structured, brief simple psychotherapeutic approach to (a) increase engagement in rewarding and adaptive activities, (b) decrease engagement in activities that maintain depression, and (c) solve problems that limit access to reward or that maintain avoidance’.^17^

Within the BASIL BA intervention, the therapist (‘BASIL Support Worker’ (BSW)) and participant worked together to develop a collaborative treatment plan to reinstate (or replace, if former activities were no longer possible because of social isolation and/or long-term conditions) behaviours that connect them to sources of positive reinforcement (valued activity). BA has the potential to address depression and loneliness in the presence of social-isolation in this way and the simplicity of BA made it suitable for delivery in the context of COVID-19.

Intervention participants were offered up to eight weekly sessions by trained BSWs, accompanied by participant materials. Participants in the intervention group were provided with a BASIL+ BA booklet. This booklet was modified to take account of UK Government guidance regarding the need for social isolation/physical distancing and enforced isolation for those people most at risk (‘clinically extremely vulnerable’ people). For example, the BASIL+ booklet discussed ways to replace activities which are no longer possible with ones that preserve social distancing whilst helping participants stay connected with the activities and people important to them; illustrative patient stories included in the booklet were modified to take account of COVID-19 restrictions. BA acknowledged the disruption to people’s lives and usual routines and encouraged the establishment of a balanced daily routine. Intervention sessions were delivered remotely via telephone. An additional offer of video call was rarely taken up. The first session was scheduled to last approximately one hour, with subsequent sessions lasting approximately 30 minutes. Where feasible and where considered appropriate and acceptable by the participant and BSW, the intervention was extended to include involvement of a participant’s informal caregiver/significant other.

**Comparator (usual GP care):** Participants in the control group received usual care as provided by their NHS and/or third sector providers. In addition, control participants were ‘signposted’ to reputable sources of self-help and information, including advice on how to keep mentally and physically well. Examples of such sources was the Public Health England (PHE) ‘Guidance for the public on the mental health and wellbeing aspects of coronavirus (COVID-19)’^18^ and Age UK.^19^

This was a pragmatic trial and no treatment was withheld by reason of participation in the BASIL+ trial in either the intervention or control arms.

### Outcome measures

Outcome measures were collected at baseline, one, three- and 12-months post-randomisation. The primary clinical outcome and endpoint was self-reported symptoms of depression, assessed by the PHQ-9,^20^ at three months. Our experience from the BASIL-C19 pilot trial^11, 12^ provided empirical evidence that most BA sessions should have been delivered by this post-randomisation point.

Other secondary outcomes were perceived loneliness (measured by the De Jong Gierveld Scale - 11 items loneliness scale, and its subscales for social loneliness (5 items) and emotional loneliness (6 items)),^21^ anxiety (measured by the GAD-7),^22^ health-related quality of life (measured by the SF-12v2 mental component scale (MCS) and physical component scale (PCS), and the EQ-5D-3L),^23^ social networks (measured by the Lubben Social Network Scale - 6 items),^24^ and questions relating to COVID-19 circumstances.

Here, we only report outcomes at one and three months since these timepoints include our primary outcome, and the 12-month outcomes are not yet available for analysis. Future analyses will also include a quantitative and economic evaluation.

### Sample size & statistical analysis

#### Sample size

The final design of the BASIL+ trial was to detect a standardised effect size of 0.3 in the primary outcome with 90% power. We drew on data from our pilot trial^11, 12^ to estimate 10% attrition (from mortality and loss to follow-up) and the correlation between baseline and primary outcome PHQ-9 score for eligible participants (0.58 amongst participants who scored ≥5 at baseline). Assuming a correlation of at least 0.5, 90% power, 5% alpha, 0.3 effect size and 10% attrition, we estimated that the BASIL+ trial needed to recruit and randomise 392 participants (power calculations performed in Stata v15). In the original trial design, we did not incorporate correlation between baseline and primary outcome and we were able to revise our study sample size to incorporate these data as the results of the BASIL pilot emerged [see supplementary file s1: Statistical Analysis Plan]. The final statistical analysis plan was approved by the independent BASIL+ Trial Steering and Data Monitoring & Ethics Committee on 15/06/2022.

#### Statistical analysis

Analyses were conducted in Stata v17 on an intention-to-treat basis using two-sided tests at the 5% significance level.^25^ The trial is reported to Consolidated Standards of Reporting Trials (CONSORT) guidelines, and the flow of participants through the trial is presented in a CONSORT diagram. Baseline data are summarised descriptively by trial arm both as randomised and as included in the primary analysis.

The primary analysis compared depression severity as measured by the PHQ-9 between the two groups using a covariance pattern mixed-effect linear regression model, incorporating data from the one and three-month follow-up time points. Treatment group, timepoint, treatment-by-time interaction, and baseline PHQ-9 score were fixed effects, with participant nested within site as random effects. An exchangeable covariance pattern for the correlation between the observations for a participant over time was specified (based on minimising the Akaike’s information criterion).^26^ Estimates of the difference in total PHQ-9 score were extracted for each timepoint as an adjusted mean difference, 95% confidence interval (CI) and p-value.

Intervention adherence, including the total number of BA sessions completed per participant and the average duration of sessions, is summarised descriptively. A Complier Average Causal Effect (CACE) analysis^27^ to assess the impact of compliance on the primary estimate was undertaken, using a two-stage instrumental variable (IV) regression approach with randomised group as the IV, adjusting for baseline score and with robust standard errors to account for clustering within site. Compliance was defined in two ways: as a continuous measure of the number of BA sessions attended; and as a dichotomous measure to indicate that at least five sessions were attended. A pre-specified subgroup analysis exploring differential intervention effects depending on baseline level of depressive symptoms (PHQ-9 score 5-9, or 10 and above) was conducted by repeating the primary analysis but including an indicator variable for whether the participant scored 5-9 or 10 or above at baseline as a covariate (rather than the continuous score) plus an interaction term between treatment allocation and baseline PHQ-9 threshold.

In light of an observed differential dropout between the trial arms, post hoc analyses were conducted to investigate missing data. To investigate the effect of missing data on the treatment effect, any baseline variables associated with non-response at the 3-month follow-up (i.e., no valid PHQ-9 score) were identified and included as covariates in the primary analysis model. Multiple imputation of the primary analysis was also conducted.

The secondary outcomes of De Jong Gierveld Loneliness Scale (two subscales separately, and total score), GAD-7, Lubben Social Network Scale, SF-12 (mental and physical health component scores separately) and EQ-5D-3L (VAS score and index value score based on the UK Tariff^28^ were analysed in a similar way to the primary outcome, swapping baseline PHQ-9 score with baseline value of the outcome as a covariate. For each of the clinical outcomes, scoring and handling of missing item level data was performed in accordance with the user guides.

Serious and non-serious adverse events are summarised by trial arm and overall.

### Ethical approval

The BASIL+ trial received ethical approval from a Research Ethics Committee (The Old Chapel, Royal Standard Place, Nottingham, NG1 6FS, UK; +44 (0)207 104 8018) on 11/12/2021 (REC Ref: 20/YH/0347). The sponsor for BASIL+ was Tees, Esk and Wear Valleys NHS Foundation Trust.

### Patient and Public Involvement (PPI)

The BASIL trial was informed by a Patient and Public Involvement Advisory Group (PPI AG) who were working with the research collective on the existing NIHR-funded research programme. This PPI AG included older adults with lived experience of mental health and/or physical health conditions, and caregivers. The PPI AG were consulted on many aspects of the trial design including (but not limited to) modification of the BA intervention for BASIL, remote recruitment of BASIL+ participants, and the relevance and readability of study recruitment information.

## Results

### Participant recruitment and characteristics

Twenty-seven general practices within England and Wales conducted database searches and mailouts. Approximately 11,900 study information packs were mailed out to potentially eligible patients between 08/02/2021 and 17/12/2021.

A total of 1325 patients expressed an interest in the study following receipt of a postal study pack, across 11 recruiting sites (entry route: permission to contact (n=1052; 79.4%), direct contact (n=194; 14.6%) or online consent (n=79; 6.0%)), of which 449 (33.9%) were identified as eligible (Figure 1). Of those who were eligible, 14 did not go on to complete a baseline assessment and were not recruited into the trial, while 435 (96.9%) were recruited and randomised into the study (n=218 to BA intervention; n=217 to usual care, with signposting). Randomisation took place from 25/02/2021 to 28/02/2022, with participants recruited across 26 general practices [one of the original 27 practices did not recruit].

**Figure 1.** consort diagram

The mean age of participants was 75.7 years (SD 6.7); 270 (62.1%) were female, and most were White [British] (n=402, 92.4%) (Table 1). Cardiovascular conditions (n=288, 66.2%) and arthritis (n=186, 42.8%) were the most frequently reported long-term health conditions. Baseline characteristics were similar between the two groups. Most participants (n=296, 68.1%) were social/physical distancing and reported adhering to UK Government’s guidance in relation to COVID-19 restrictions “all of the time” (n=312, 71.7%) at baseline. Nearly half (n=200, 46.0%) reported living alone, and the majority (n=364, 83.7%) had received one or both doses of a COVID vaccine at the point of trial entry. See supplementary file s2.

**Table 1:** Baseline demographics of participants as randomised and as analysed

|  | As randomised |  |  | As analysed |  |  |
| --- | --- | --- | --- | --- | --- | --- |
|  | Intervention<br>(n=218) | Usual Care<br>(n=217) | Total<br>(n=435) | Intervention<br>(n=181) | Usual Care<br>(n=203) | Total<br>(n=384) |
| **Age, years** |  |  |  |  |  |  |
| Mean (SD) | 75.2 (6.4) | 76.2 (6.9) | 75.7 (6.7) | 75.0 (6.2) | 75.9 (6.8) | 75.5 (6.5) |
| **Gender, n (%)** |  |  |  |  |  |  |
| Male | 81 (37.2) | 84 (38.7) | 165 (37.9) | 60 (33.1) | 76 (37.4) | 136 (35.4) |
| Female | 137 (62.8) | 133 (61.3) | 270 (62.1) | 121 (66.9) | 127 (62.6) | 248 (64.6) |
| **LTC<sup>1</sup>, n (%)** |  |  |  |  |  |  |
| Cardiovascular Conditions | 144 (66.1) | 144 (66.4) | 288 (66.2) | 115 (63.5) | 136 (67.0) | 251 (65.4) |
| Arthritis | 98 (45.0) | 88 (40.6) | 186 (42.8) | 87 (48.1) | 82 (40.4) | 169 (44.0) |
| Diabetes | 77 (35.3) | 60 (27.6) | 137 (31.5) | 62 (34.3) | 57 (28.1) | 119 (31.0) |
| Respiratory Conditions | 60 (27.5) | 61 (28.1) | 121 (27.8) | 52 (28.7) | 56 (27.6) | 108 (28.1) |
| Chronic Pain | 44 (20.2) | 37 (17.1) | 81 (18.6) | 38 (21.0) | 35 (17.2) | 73 (19.0) |
| Cancer | 26 (11.9) | 26 (12.0) | 52 (12.0) | 21 (11.6) | 21 (10.3) | 42 (10.9) |
| Neurological Conditions | 23 (10.6) | 16 (7.4) | 39 (9.0) | 22 (12.2) | 15 (7.4) | 37 (9.6) |
| Osteoporosis | 19 (8.7) | 14 (6.5) | 33 (7.6) | 18 (9.9) | 12 (5.9) | 30 (7.8) |
| Stroke | 12 (5.5) | 16 (7.4) | 28 (6.4) | 7 (3.9) | 15 (7.4) | 22 (5.7) |
| Other<sup>2</sup> | 46 (21.1) | 51 (23.5) | 97 (22.3) | 38 (21.0) | 47 (23.2) | 85 (22.1) |
| **Ethnicity, n (%)** |  |  |  |  |  |  |
| White (English/Welsh/Scottish/N. Irish/British) | 201 (92.2) | 201 (92.6) | 402 (92.4) | 169 (93.4) | 188 (92.6) | 357 (93.0) |
| White (Irish) | 3 (1.4) | 2 (0.9) | 5 (1.1) | 3 (1.7) | 2 (1.0) | 5 (1.3) |
| White (Other white background) | 5 (2.3) | 6 (2.8) | 11 (2.5) | 3 (1.7) | 6 (3.0) | 9 (2.3) |
| Asian (Indian) | 1 (0.5) | 1 (0.5) | 2 (0.5) | 1 (0.6) | 1 (0.5) | 2 (0.5) |
| Asian (Pakistani) | 0 (0.0) | 1 (0.5) | 1 (0.2) | 0 (0.0) | 1 (0.5) | 1 (0.3) |
| Asian (Chinese) | 1 (0.5) | 0 (0.0) | 1 (0.2) |  |  |  |
| Asian (Other) | 2 (0.9) | 1 (0.5) | 3 (0.7) | 2 (1.1) | 1 (0.5) | 3 (0.8) |
| Black (African) | 2 (0.9) | 4 (1.8) | 6 (1.4) | 0 (0.0) | 3 (1.5) | 3 (0.8) |
| Black (Caribbean) | 1 (0.5) | 1 (0.5) | 2 (0.5) | 1 (0.6) | 1 (0.5) | 2 (0.5) |
| Prefer not to say | 1 (0.5) | 0 (0.0) | 1 (0.2) | 1 (0.6) | 0 (0.0) | 1 (0.3) |
| Mixed/Multiple Ethnic Groups (Other) | 1 (0.5) | 0 (0.0) | 1 (0.2) | 1 (0.6) | 0 (0.0) | 1 (0.3) |
| **Smoking Status, n (%)** |  |  |  |  |  |  |
| I have never smoked | 83 (38.1) | 93 (42.9) | 176 (40.5) | 67 (37.0) | 86 (42.4) | 153 (39.8) |
| I currently smoke | 20 (9.2) | 16 (7.4) | 36 (8.3) | 16 (8.8) | 14 (6.9) | 30 (7.8) |
| I am an ex-smoker | 115 (52.8) | 108 (49.8) | 223 (51.3) | 98 (54.1) | 103 (50.7) | 201 (52.3) |
| **Alcohol Intake (2+ units daily), n (%)** |  |  |  |  |  |  |
| Yes | 36 (16.5) | 39 (18.0) | 75 (17.2) | 30 (16.6) | 36 (17.7) | 66 (17.2) |
| No | 180 (82.6) | 176 (81.1) | 356 (81.8) | 149 (82.3) | 165 (81.3) | 314 (81.8) |
| Don't know | 2 (0.9) | 2 (0.9) | 4 (0.9) | 2 (1.1) | 2 (1.0) | 4 (1.0) |
| **Education after school leaving age, n (%)** |  |  |  |  |  |  |
| Yes | 136 (62.4) | 122 (56.2) | 258 (59.3) | 116 (64.1) | 113 (55.7) | 229 (59.6) |
| **Marital Status, n (%)** |  |  |  |  |  |  |
| Married | 95 (43.6) | 104 (47.9) | 199 (45.7) | 76 (42.0) | 100 (49.3) | 176 (45.8) |
| Widowed | 57 (26.1) | 64 (29.5) | 121 (27.8) | 50 (27.6) | 60 (29.6) | 110 (28.6) |
| Divorced/Separated | 37 (17.0) | 32 (14.7) | 69 (15.9) | 31 (17.1) | 27 (13.3) | 58 (15.1) |
| Single | 16 (7.3) | 14 (6.5) | 30 (6.9) | 15 (8.3) | 13 (6.4) | 28 (7.3) |
| Cohabiting | 8 (3.7) | 2 (0.9) | 10 (2.3) | 7 (3.9) | 2 (1.0) | 9 (2.3) |
| Civil Partnership | 4 (1.8) | 1 (0.5) | 5 (1.1) | 2 (1.1) | 1 (0.5) | 3 (0.8) |
| Not reported | 1 (0.5) | 0 (0.0) | 1 (0.2) | 0 (0.0) | 0 (0.0) | 0 (0.0) |

<sup>1</sup>Long-term health conditions were participant self-reported and were not mutually exclusive.
<sup>2</sup>Other long-term health conditions included: hypothyroidism (n=35); gastrointestinal conditions (n=19); renal conditions (n=13); osteoarthritis (n=6); mental health conditions (n=5); inflammatory conditions (n=5); dermatological conditions (n=5); sleep apnoea (n=4); sensory conditions (n=4); haematological conditions (n=4); pain related conditions (n=3); unknown (n=3); low immunity conditions (n=2); B12 deficiency (n=2); gynaecological conditions (n=1); hyperthyroidism (n=1); long COVID (n=1); lymphoedema (n=1); autoimmune conditions (n=1). Participants may have reported multiple 'other' conditions.

### Engagement with the BASIL intervention

Intervention participants attended an average of 5.2 (of eight) sessions (SD 2.9), 63.8% (n=139) attended at least 5 sessions and just over a third (n=80, 36.7%) attended all eight. BA sessions took place over an average of 7.9 weeks (SD 3.7). On average, the first session took place 15.4 days after randomisation (SD 10.9, median 13). From data across 1,114 sessions, sessions lasted on average 36.2 minutes (SD 13.2).

### Participant Follow Up and Retention

Overall, 358 (82.3%) participants (n=168, 77.1% intervention, n=190, 87.6% usual care) completed the one-month follow-up questionnaire and 359 (82.5%) participants (n=166, 76.1% intervention, n=193, 88.9% usual care) completed the three-month follow-up questionnaire (Table 2). Responses were higher in the usual care arm at both timepoints.

**Table 2:** Response rates at one and three months by trial arm.

|  | **Intervention**<br>**(n=218)** | **Usual Care**<br>**(n=217)** | **Total**<br>**(n=435)** |
| --- | --- | --- | --- |
| **1-month, n (%)** | 168 (77.1) | 190 (87.6) | 358 (82.3) |
| **3-months, n (%)** | 166 (76.1) | 193 (88.9) | 359 (82.5) |

### Outcome data and between group comparisons at 1 and 3 months [primary outcome point]

Of the 435 participants randomised, 384 (88.3%) were included in the primary analysis; 51 (11.7%) were omitted due to non-completion of both the one and three-month follow up questionnaires. No participants were excluded from the primary analysis on the basis of missing baseline covariates (PHQ-9 score at baseline), as this was provided for all randomised participants.

Patient reported outcome measures (unadjusted) and between group comparisons (adjusted) are provided by group in Tables 3 and 4. On average, PHQ-9 scores decreased over time in both groups (overall mean: baseline 9.8 (SD 4.3), one-month 7.7 (SD 5.0); three-month 6.4 (SD 4.8)). The observed correlation between baseline and outcome PHQ-9 scores was 0.48 (95% CI 0.40 to 0.56) at one-month and 0.42 (95% CI 0.33 to 0.50) at three months. There was evidence of a difference in depression severity, favouring the intervention group across the follow up period; at both one-month (PHQ-9 score adjusted mean difference -1.25, 95% CI -2.15 to -0.35, p=0.01) and three months (primary endpoint of primary outcome, adjusted mean difference -1.65, 95% CI -2.54 to - 0.75, p<0.001), see Figure 2 and Table 4. Findings were robust to sensitivity analyses.

**Figure 2:**
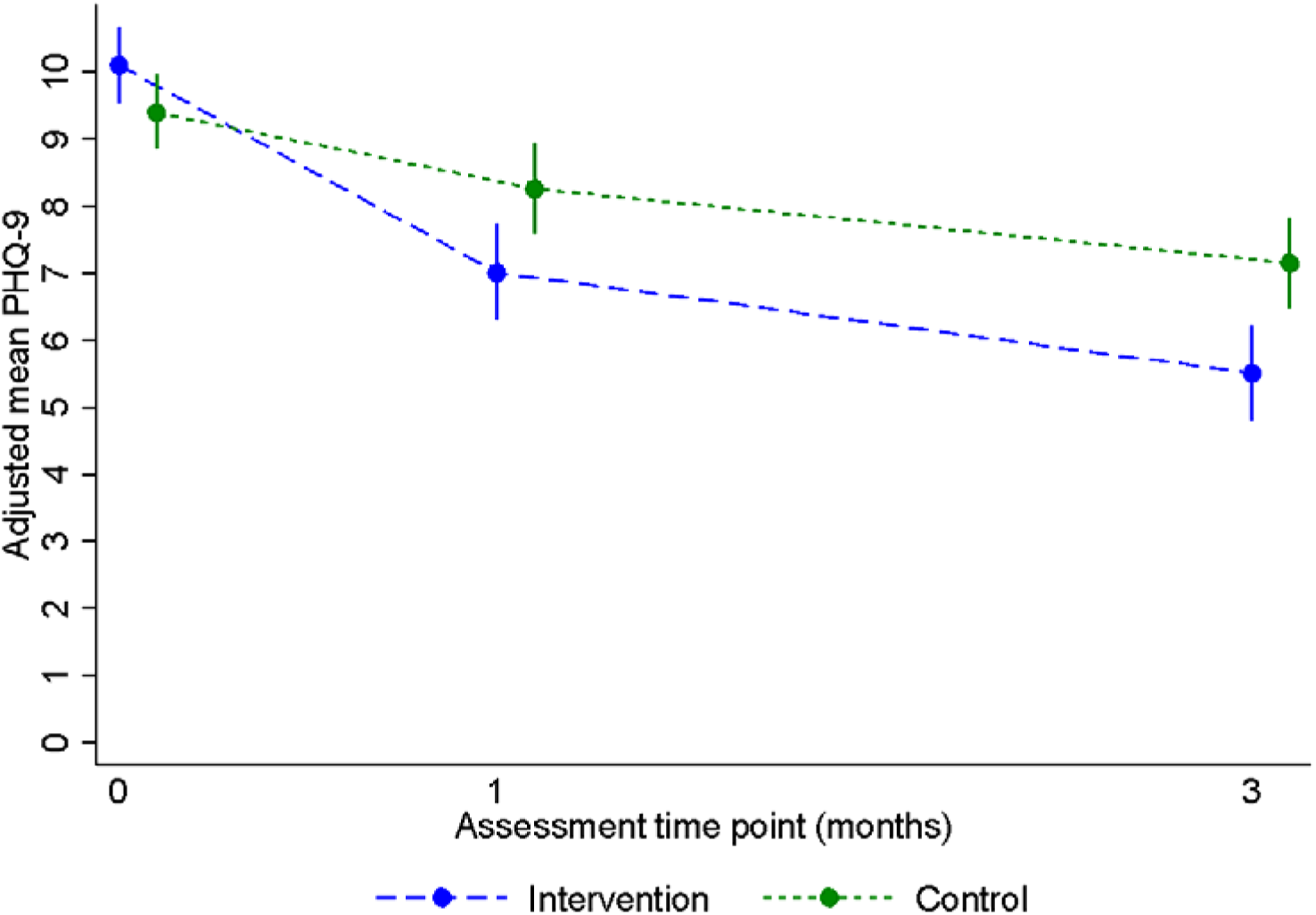
PHQ-9 adjusted depression severity across the follow up period [1 and 3 months]

**Table 3:** Raw summaries of patient Reported Outcome Measures by trial arm and timepoint

| Outcome Measure | Intervention |  | Usual Care |  |
| --- | --- | --- | --- | --- |
|  | n | Mean (SD) | n | Mean (SD) |
| **PHQ-9** |  |  |  |  |
| Baseline | 218 | 10.1 (4.3) | 217 | 9.4 (4.2) |
| 1-month | 167 | 7.2 (4.5) | 190 | 8.1 (5.5) |
| 3-month | 165 | 5.7 (4.0) | 192 | 6.9 (5.3) |
| **GAD-7** |  |  |  |  |
| Baseline | 218 | 5.7 (4.4) | 216 | 6.2 (4.9) |
| 1-month | 164 | 4.7 (4.2) | 189 | 5.7 (4.7) |
| 3-month | 164 | 3.8 (3.8) | 192 | 4.6 (4.6) |
| **De Jong Gierveld Loneliness Scale** |  |  |  |  |
| Baseline | 218 | 5.5 (3.1) | 217 | 5.7 (3.1) |
| 1-month | 164 | 5.0 (3.3) | 189 | 5.1 (3.2) |
| 3-month | 162 | 4.6 (3.1) | 190 | 4.9 (3.3) |
| **De Jong Gierveld Emotional Loneliness Subscale** |  |  |  |  |
| Baseline | 218 | 3.3 (1.8) | 217 | 3.5 (1.8) |
| 1-month | 164 | 2.9 (1.9) | 189 | 3.0 (1.8) |
| 3-month | 162 | 2.6 (1.9) | 190 | 3.0 (1.9) |
| **De Jong Gierveld Social Loneliness Subscale** |  |  |  |  |
| Baseline | 218 | 2.2 (1.8) | 217 | 2.2 (1.8) |
| 1-month | 164 | 2.0 (1.8) | 189 | 2.1 (1.9) |
| 3-month | 162 | 2.0 (1.8) | 190 | 1.9 (1.9) |
| **LSNS6** |  |  |  |  |
| Baseline | 218 | 13.8 (5.6) | 217 | 13.2 (5.8) |
| 1-month | 164 | 13.9 (6.1) | 189 | 13.7 (6.1) |
| 3-month | 161 | 14.7 (6.0) | 190 | 14.3 (6.4) |
| **SF-12v2 (Physical Health Component Score)** |  |  |  |  |
| Baseline | 218 | 37.3 (11.3) | 217 | 37.6 (10.8) |
| 1-month | 164 | 37.0 (11.1) | 189 | 37.7 (11.6) |
| 3-month | 161 | 37.6 (11.8) | 190 | 38.1 (11.0) |
| **SF-12v2 (Mental Health Component Score)** |  |  |  |  |
| Baseline | 218 | 44.4 (10.0) | 217 | 44.0 (10.2) |
| 1-month | 164 | 46.1 (9.8) | 189 | 45.8 (10.6) |
| 3-month | 161 | 48.3 (9.3) | 190 | 46.4 (10.8) |
| **EQ-5D-3L index value score** |  |  |  |  |
| Baseline | 217 | 0.608<br>(0.267) | 217 | 0.619<br>(0.279) |
| 1-month | 164 | 0.631<br>(0.277) | 187 | 0.622<br>(0.284) |
| 3-month | 161 | 0.642<br>(0.275) | 190 | 0.642<br>(0.276) |
| **EQ-5D-3L VAS** |  |  |  |  |
| Baseline | 218 | 58.8 (20.2) | 216 | 59.3 (19.2) |
| 1-month | 164 | 61.9 (19.4) | 187 | 59.7 (20.9) |
| 3-month | 160 | 61.3 (20.1) | 190 | 60.2 (22.4) |

PHQ, Patient Health Questionnaire; GAD, Generalised Anxiety Disorder; SF, Short Form.
Range of possible scores: PHQ-9 0-27; GAD-7 0-21; De Jong Gierveld total 0-11 – Emotional subscale 0-6, Social subscale 0-5 (higher score indicates worse outcome). SF-12v2 subscales 0-100; EQ-5D-3L index value score -0.285 (worse than death) -1 (full health); EQ-5D-3L VAS 0-100 (higher score indicates better outcome).

**Table 4.** Adjusted mean differences between the BA and usual care groups by time point

|  | Adjusted<sup>1</sup> mean difference between groups at for primary and secondary outcomes |  |  |  |
| --- | --- | --- | --- | --- |
|  | Intervention Mean (95% CI) | Usual Care Mean (95% CI) | Difference (95% CI) | p-value |
| **Primary Outcome** |  |  |  |  |
| PHQ-9 (3-months) | 5.5 (4.8, 6.2) | 7.2 (6.5, 7.8) | -1.65 (-2.54, -0.75) | <0.001 |
| **Sensitivity analyses of PHQ-9** |  |  |  |  |
| *Adjusting for predictors of missing data<sup>2</sup>* |  |  |  |  |
| 1-month | 7.1 (6.3, 7.8) | 8.2 (7.5, 8.9) | -1.18 (-2.06, -0.29) | 0.01 |
| 3-months | 5.5 (4.8, 6.2) | 7.1 (6.4, 7.8) | -1.58 (-2.47, -0.69) | 0.00 |
| *Analysis using multiply imputed data<sup>3</sup>* |  |  |  |  |
| 1-month | 7.1 (6.4, 7.8) | 8.3 (7.7, 9.0) | -1.19 (-2.10, -0.29) | 0.01 |
| 3-months | 5.6 (4.9, 6.3) | 7.2 (6.6, 7.8) | -1.63 (-2.50, -0.77) | <0.001 |
| **Secondary Outcomes at 1-month** |  |  |  |  |
| PHQ-9 | 7.0 (6.3, 7.7) | 8.3 (7.6, 8.9) | -1.25 (-2.15, -0.35) | 0.01 |
| GAD-7 | 4.9 (4.3, 5.4) | 5.5 (5.0, 6.0) | -0.65 (-1.41, 0.11) | 0.09 |
| De Jong Gierveld Loneliness Scale | 5.1 (4.7, 5.5) | 5.1 (4.7, 5.5) | -0.03 (-0.49, 0.43) | 0.89 |
| De Jong Gierveld Emotional Loneliness Scale | 3.0 (2.8, 3.3) | 3.0 (2.8, 3.2) | 0.05 (-0.24, 0.33) | 0.75 |
| De Jong Gierveld Social Loneliness Scale | 2.0 (1.8, 2.3) | 2.14 (1.9, 2.4) | -0.09 (-0.37, 0.19) | 0.53 |
| Lubben Social Network Scale (6-items) | 13.5 (12.7, 14.2) | 14.2 (13.5, 14.9) | -0.72 (-1.57, 0.14) | 0.10 |
| SF-12v2 (Physical Health Component Score) | 37.1 (36.1, 38.2) | 37.9 (36.9, 38.9) | -0.80 (-2.24, 0.68) | 0.30 |
| SF-12v2 (Mental Health Component Score) | 46.0 (44.7, 47.3) | 45.9 (44.7, 47.1) | 0.10 (-1.66, 1.87) | 0.90 |
| EQ-5D-3L index value score | 0.639 (0.599, 0.679) | 0.623 (0.585, 0.661) | 0.016 (-0.031, 0.063) | 0.51 |
| EQ-5D-3L VAS score | 61.7 (59.0, 64.4) | 60.1 (57.6, 62.6) | 1.64 (-2.03, 5.30) | 0.38 |
| **Secondary Outcomes at 3-months** |  |  |  |  |
| GAD-7 | 3.9 (3.3, 4.5) | 4.6 (4.0, 5.1) | -0.67 (-1.43, 0.09) | 0.08 |
| De Jong Gierveld Loneliness Scale | 4.6 (4.2, 5.0) | 5.0 (4.6, 5.4) | -0.42 (-0.88, 0.04) | 0.08 |
| De Jong Gierveld Emotional Loneliness Scale | 2.6 (2.4, 2.9) | 3.0 (2.8, 3.2) | -0.37 (-0.68, -0.06) | 0.02 |
| De Jong Gierveld Social Loneliness Scale | 1.9 (1.7, 2.2) | 1.99 (1.7, 2.3) | -0.05 (-0.33, 0.23) | 0.72 |
| Lubben Social Network Scale (6-items) | 14.3 (13.5, 15.0) | 14.6 (13.9, 15.3) | -0.36 (-1.21, 0.50) | 0.41 |
| SF-12v2 (Physical Health Component Score) | 37.4 (36.2, 38.6) | 37.9 (36.8, 39.0) | -0.50 (-2.14, 1.10) | 0.53 |
| SF-12v2 (Mental Health Component Score) | 48.4 (47.1, 49.7) | 46.4 (45.2, 47.6) | 1.99 (0.22, 3.76) | 0.03 |
| EQ-5D-3L index value score | 0.633<br>(0.593, 0.673) | 0.641<br>(0.604, 0.679) | -0.008<br>(-0.055, 0.038) | 0.73 |
| EQ-5D-3L VAS score | 60.8 (58.1, 63.5) | 60.0 (57.5, 62.4) | 0.84 (-2.83, 4.52) | 0.65 |

<sup>1</sup>Adjusted for baseline measure of the outcome as a covariate; <sup>2</sup>As primary analysis model additionally adjusting for gender, educated to degree level or equivalent, baseline total De Jong Gierveld Loneliness Scale score and baseline Lubben Social Network Scale score; <sup>3</sup>As primary analysis model, using data derived by multiple imputation based on predictors: allocation, gender, age, ethnicity, number of comorbidities, degree, marital status and baseline scores for PHQ-9 and all secondary outcomes.

For the measure of loneliness (De Jong Gierveld Loneliness Score) there was evidence of a difference in emotional loneliness, favouring the intervention group, at three months (adjusted mean difference -0.37, 95% CI -0.68 to -0.06, p=0.02), but there was no evidence of a difference for social loneliness (-0.05, 95% CI -0.33, 0.23, p=0.72). For the total Loneliness Score, the adjusted mean difference indicated a lower severity of loneliness at three months in the intervention group (-0.4, 95% CI -0.88 to 0.04, p=0.08), but the 95% CI contained zero. At one-month there was no evidence of a statistically significant benefit in any aspect of measured loneliness [emotional loneliness p=0.75, social loneliness p=0.53, total loneliness p=0.89].

There was a signal of reduced anxiety (GAD-7) in the intervention group at one-month (adjusted mean difference -0.65, 95% CI -1.41 to 0.11, p=0.09) and three months (adjusted mean difference - 0.67, 95% CI -1.43 to 0.09, p=0.08), but the 95% CIs contained zero.

In the measure of social networks (Lubben Social Network Scale), there was no evidence of a statistically significant difference at one-month (adjusted mean difference -0.72, 95% CI -1.57 to 0.14, p=0.10) or at three months (adjusted mean difference -0.36, 95% CI -1.21 to 0.50, p=0.41).

There was no evidence of a statistically significant difference in health-related quality of life as measured by the EQ-5D-3L index or VAS scores between treatment groups at either one or three months (Table 4).

### Compliance Engagement with the BASIL Intervention

In the CACE analysis defining compliance as a continuous measure, there was an indication that for every BA session attended there was a reduction in depression severity of 0.26 points on the PHQ-9 at three months (95% CI -0.42 to -0.10, p=0.002). Where compliance was treated as a dichotomous measure, there was an indication of a reduction in depression severity of two points on the PHQ-9 at three months when at least five BA sessions were completed (2.02, 95% CI -3.29 to -0.76, p=0.002).

### Subgroup Analysis

Of the 435 randomised participants, 251 (57.7%) scored 5-9 on the PHQ-9 at baseline (n=115, 52.8% intervention, n=136, 62.7% usual care), and 184 (42.3%) scored ten or more (n=103, 47.2% intervention, n=81, 37.3% usual care). There was weak evidence of an interaction between PHQ-9 score at baseline and trial arm (interaction effect p=0.10).

The adjusted mean difference between the intervention and usual care groups was estimated at -1.13 (95% CI –2.26 to 0.01, p=0.05) among those scoring 5-9 on the PHQ-9 at baseline, but a larger effect was seen amongst those scoring 10 or more at baseline (-2.48, 95% CI -3.81 to 1.16, p<0.001).

### BASIL+ Living systematic review

When we added the results of the BASIL+ trial to a pre-registered living systematic review [PROSPERO CRD42021298788],^12^ we found strong evidence of effect on depression outcomes [random effects pooled standardised mean difference -0.32, 95% CI -0.48 to -0.16, 10 studies, n=1,210 participants] and also loneliness [random effects pooled standardised mean difference -0.44, 95% CI -0.64 to -0.24, 13 studies, n=1,421 participants] in the short-term (<6 months). See Figure 3 & research in context panel.

**Figure 3:**
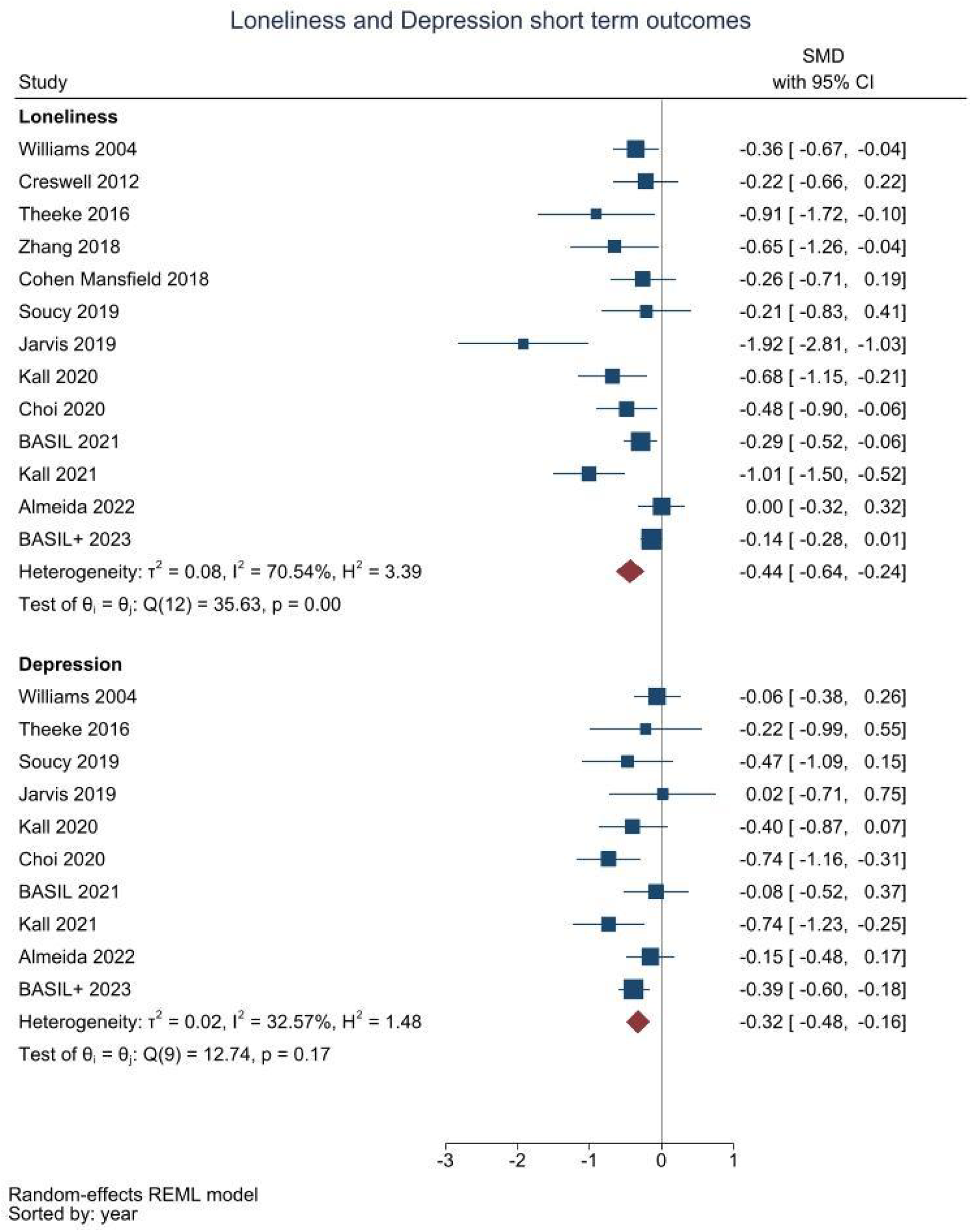
Random effects meta-analysis from a living systematic review of cognitive and/or behavioural trials for depression and loneliness, incorporating the results of the BASIL+ trial

## Panel: Research in context

### Evidence before this study

Before designing the BASIL+ trial we updated reviews of Behavioural Activation. We searched key databases (MEDLINE; EMBASE; CINAHL; PsycINFO; HMIC; CENTRAL and DARE, English language from inception to June 2020) for Collaborative Care studies and trials of Behavioural Activation, and independently extracted data. We included randomised controlled trials conducted in any country or care setting with adult and older adults with depression. There were no large-scale UK trials of Collaborative Care and no large-scale trials of Behavioural Activation [and no meta-analyses of trials] addressing loneliness in socially isolated older people in any setting.

### Added value of this study

The BASIL trial demonstrates that it is feasible to deliver a BA intervention for older people with long-term conditions who were socially isolated in response to COVID-19. Older people readily engaged with behavioural activation as a remote (telephone)-delivered psychological intervention. We found significant non-chance reductions in both depression and loneliness at 3 months.

We also planned to assimilate the results of the BASIL+ trial into a living systematic review [pre-registered PROSPERO CRD42021298788]. There is now clear evidence of benefit for depression outcomes [random effects pooled standardised mean difference -0.32, 95% CI -0.48 to -0.16, 10 studies, n=1,210 participants] and also loneliness [random effects pooled standardised mean difference -0.44, 95% CI -0.64 to -0.24, 13 studies, n=1,421 participants] in the short-term (<6 months). BASIL+ contributes to the trial-based evidence for behavioural and/or cognitive interventions, and is the largest trial in this area to date.

### Implications of all the available evidence

Behavioural activation was a plausible intervention to mitigate depression and loneliness in at risk older populations during the COVID pandemic, and this evidence will be useful to practitioners and policy makers beyond the pandemic in preventing loneliness in vulnerable populations.

## Discussion

We conducted an RCT within primary care of a psychosocial intervention involving a large number of geographically diverse sites (including the NHS and Age UK organisations) in the later stages of the COVID-19 pandemic for older people with long-term conditions. This population was vulnerable to the psychological impacts of COVID-19 restrictions. Our main finding is that Behavioural Activation, a brief telephone-delivered psychological intervention adapted to mitigate depression and loneliness, showed evidence of short-term positive psychological impact. There was evidence of an immediate benefit in terms of depression severity at one month, and this was evident at the primary trial endpoint of three months. For important secondary outcome measures there was strong evidence that BA reduced levels of emotional loneliness (but not social loneliness) and also improved quality of life relating to mental health (but not physical health). There was some evidence that BA reduced levels of anxiety. Psychological benefit was greatest for those with more severe depression and among people who engaged with 5 or more BA sessions.

The BASIL+ trial followed a developmental phase where BA was first adapted to meet the needs of socially isolated older adults at the beginning of the COVID-19 pandemic, and was tested in an external pilot trial.^11^. The BASIL+ trial was co-produced and supported by a specially convened COVID-19 research prioritisation and delivery mechanism, focussed mainly (though not exclusively) on the roll out of randomised trials in the response to the COVID pandemic. The BASIL+ trial was one of only two trials of psychosocial interventions to address unmet psychological needs during this unprecedented time. The BASIL+ trial represents an advancement in trials-based evidence in this under-researched area. The BASIL+ trial triangulates with existing research into behavioural activation to reduce and prevent depression in vulnerable populations, (including older people^13, 29^) and BASIL+ extends this research knowledge base to vulnerable populations during pandemic conditions. The BASIL+ trial was also designed to mitigate levels of loneliness in a vulnerable population, and a novel finding from BASIL+ is that a scalable behavioural intervention reduces levels of emotional loneliness. Trials of psychosocial intervention targeting loneliness, to date, have been small scale and the BASIL+ trial is, by some margin, the largest trial of a behavioural intervention ever conducted to mitigate loneliness. The need for scalable psychological solutions to social isolation and loneliness was identified as an important research priority at the commencement of the pandemic,^7^ and the BASIL+ trial closes this evidential gap.

There were limitations in the BASIL+ trial. The first limitation we highlight is that there was differential attrition. Whilst follow up rates were high (and in line with predicted rates of retention based on our pilot), we noted some differential rates of attrition between intervention and control arms. However, findings for the primary outcome were robust to post-hoc sensitivity analyses investigating the effect of missing data. The second limitation is that we were unable to blind participants to trial allocation and we sought to mitigate this potential bias by ensuring that, where possible, the assessment of outcome was undertaken by researchers who were blinded to treatment allocation. A further limitation is that our original proposed sample size was not feasible during the trial recruitment period. However, we were able to recruit slightly above the revised sample size target which was calculated to account for correlation between baseline and follow-up measurements; though the observed correlation between baseline and month three PHQ-9 score was slightly lower than the one we assumed (0.42 vs 0.50). Finally, we designed a pragmatic trial to assess effectiveness of a behavioural intervention, but the choice of comparator of usual care does not allow the specific effects of behavioural approaches to be assumed. BASIL+ was designed as a two-arm trial with a ‘usual care plus signposting’ comparator, and an alternative design might have been to add a third arm with an attention control condition. Our final design was guided by the BASIL+ lived experience PPI expert group and the optimum design was informed by theory and evidence of ‘what works’ from prior research.^30^ We note that our control condition included ‘signposting’ to ensure that participants were also offered a credible self-help option in line with best practice and policy recommendations across the pandemic. We also note a ‘dose response’ effect from our CACE analysis and this supports the principle that engagement with the intervention maximised the effectiveness of behavioural activation.

The COVID-19 pandemic prompted a number of studies to understand the impacts of COVID-19,^31^ but there have been very few studies to evaluate psychosocial interventions to mitigate psychological impact. ^8^ Beyond the pandemic a clinical priority and policy imperative is to identify a brief and scalable intervention to prevent and mitigate loneliness, particularly in older people ^32, 33^. The BASIL trials programme will be informative in improving the mental health of populations in socially isolated at-risk populations after the pandemic has passed. ^6^

Loneliness is a clinically-, socially- and economically-important phenomenon, that is now increasingly recognised as a threat to population health. There are several approaches that can be helpful to policy makers, but also important evidential gaps in terms of ‘what works’ in preventing or mitigating loneliness.^34^ This is a rapidly advancing area and the successful delivery of the BASIL+ trial during pandemic conditions contributes to an evolving evidence base in response to social isolation and the risk of psychological deterioration in at-risk groups. Looking to the future BA can be used to mitigate depression and risk of loneliness in the presence of shocks to health systems and populations, such as future pandemics or other shocks that increase anxiety and depression among vulnerable groups, such as the climate emergency.^33^

## Contributions of the authors

SG, DE, CCG, EL, DMcM, CH, DB and SB planned the trial, contributed to the trial design and drafted the trial protocol. SG and DE led manuscript writing, managed the trial as chief investigators, and critically revised the manuscript. SG, DE, EL, DMcM, CCG, CH, PC, GTT, AC, TG, AHi, KL, SDS, CS, EN, TO, HW and JW contributed to trial design and trial management meetings.

SG, CCG, DE, DMcM, DB and CS designed and contributed to the intervention and BSW training materials, and DB, DMcM, CCG and DE delivered the BSW training. EL led the day-to-day management of the trial, and SB, LB, LA and RW were the trial coordinators. DB, SC and DMcM provided BSW clinical supervision. SB, LB, AH, ER, LS, LA and RW facilitated participant recruitment and follow-up data collection, and participated in trial management meetings. ER and LS assisted with BA intervention delivery. CF, KB and CH developed the statistical analysis plan and analysed the quantitative data. All authors contributed to the drafts of manuscripts and read the final manuscript. The York Trials Unit acts as data custodians.

## Supporting information

Statistical Analysis Plan and protocol

## Data Availability

Anonymised data will be made available upon reasonable request, which must include a protocol and statistical analysis plan and not be in conflict with our prespecified publication plan, consistent with our data sharing policy (available on request from SG). The BASIL research collective is especially keen that the BASIL data contributes to prospective meta-analyses and individual patient data meta-analyses. Requests for data sharing will be considered by SG and the independent trial steering and data monitoring committee.

## Acknowledgements

We would like to thank: the participants for taking part in the trial, general practices and North East and North Cumbria Local Clinical Research Network staff for identifying and facilitating recruitment of participants, participating NHS Trusts and Age UK organisations for recruitment, intervention delivery and follow ups, the independent Joint Programme/Trial Steering and Data Monitoring and Ethics Committee members for overseeing the study, and our PPI AG members for their insightful contributions and collaboration.

