## Supplementary material for "Mitigating the psychological impacts of COVID-19 restrictions on older people: The UK Behavioural Activation in Social Isolation (BASIL+) COVID-19 Urgent Public Health (UPH) trial and living systematic review": Statistical Analysis Plan and protocol

### BASIL+

#### Behavioural Activation in Social IsoLation (BASIL+)

STATISTICAL ANALYSIS PLAN

Draft v1.1

**York Trials Unit**
Department of Health Sciences
University of York
York, Y010 5DD

**Version date:** 26/09/2022
**Authors:** Kalpita Baird, Caroline Fairhurst and Prof. Catherine Hewitt
**Chief Investigator:** Prof. David Ekers and Prof. Simon Gilbody
**Trial Manager/Coordinator:** Dr Liz Littlewood and Lauren Burke

#### 1. Document scope and relevant SOPs and guidance documents

This statistical analysis plan (SAP) deals only with the statistical analysis of clinical effectiveness; the cost-effectiveness analysis will be detailed in a separate plan. This SAP was written following completion of recruitment but prior to the end of primary outcome follow-up and database lock. The SAP was prepared according to York Trials Unit (YTU) standard operating procedures and guidance documents.

#### 2. Definition of terms/acronyms

A definition of any terms or acronyms used in the SAP is provided in this section.

| | |
|---|---|
| AE | Adverse Event |
| BA | Behavioural Activation |
| BASIL | Behavioural Activation in Social IsoLation |
| BSW | BASIL+ Support Worker |
| CACE | Complier Average Causal Effect |
| CONSORT | Consolidated standards of reporting trials |
| CRF | Case Report Form |
| DASS | Depression Anxiety Stress Scale |
| GAD | Generalised Anxiety Disorder |
| LTC | Long Term Condition |
| RCT | Randomised Controlled Trial |
| PHQ | Patient Health Questionnaire |
| PIS | Participant Information Sheet |
| SAE | Serious Adverse Event |
| SAP | Statistical Analysis Plan |
| SWAT | Study Within a Trial |
| YTU | York Trials Unit |

#### 3. Design

BASIL+ is a pragmatic, multi-centre, two-arm, parallel group randomised controlled trial (RCT) with allocation at the individual patient level. Eligible patients (older adults aged 65 years and over, with two or more physical long term conditions (LTCs) or a condition that may indicate they are within a ‘clinically extremely vulnerable’ group in relation to COVID-19 [1], and a score of five or higher on the Patient Health Questionnaire (PHQ-9) [2]; putting them at risk of clinical depression or indicating already-established depressive symptoms) will be enrolled into the trial. Behavioural Activation (BA) is a practical psychological treatment that explores how physical inactivity and low mood are linked and aims to help people maintain or introduce activities that are important to them. Participants will be randomised to one of two arms: Behavioural Activation (BA), or usual care with signposting information.

An external pilot RCT (BASIL-C19) was conducted (three-month follow-up concluded in April 2021, and 12-month in December 2021) to test the feasibility of the BASIL intervention and remote trial delivery [including recruitment and follow-up]. The pilot RCT indicated that BA was a plausible intervention to mitigate the psychological impacts of COVID-19 isolation for older adults, is acceptable and can be feasibly delivered remotely and at scale [3].

The BASIL+ trial also includes an embedded Study Within a Trial (SWAT) to evaluate the effectiveness of a one-page infographic alongside the Participant Information Sheet (PIS) vs PIS only, on participant recruitment and response rates. Analyses of this SWAT are detailed in this SAP.

Full details of the background and design of the BASIL+ trial are presented in the published protocol [3].

#### 4. Trial Objectives

##### 4.1 Primary objective

The overarching aim of the BASIL trials programme of research is to rapidly develop a feasible, acceptable, clinical, and cost-effective brief psychological intervention (BA) to improve depression and loneliness in older adults with physical-mental multi-morbidity during isolation.

The primary objective is to undertake a parallel group RCT to test the hypothesis that BA is superior to usual care [with signposting information] in preventing and mitigating depression and loneliness among older people with multimorbidity during isolation.

##### 4.2 Secondary objectives

Further objectives include:

a. To establish the cost effectiveness of BA in preventing and mitigating depression and loneliness among older people with multimorbidity during isolation (the analysis of which will be detailed in a separate Health Economic Analysis Plan and not discussed further here).
b. Assess acceptability of, and plan the implementation for, BA for older people with multimorbidity during isolation (details of this process/qualitative evaluation will be provided separately).

#### 5. Follow-up

Participants will be asked to complete participant self-reported questionnaires at one, three- and 12-months post-randomisation (Table 1).

In addition, participants in the intervention group will be offered up to eight BA sessions over a 12-week period, delivered by a trained BASIL Support Worker (BSW) and supported by a self-help booklet. Symptom monitoring at each intervention session will be undertaken using the depression scale of the Depression Anxiety Stress Scale (DASS). Scores on the DASS will be used to guide decision making by BSWs, guided by supervision provided by clinical members of the BASIL+ study team.

*Table 1: BASIL+ Data Collection Schedule*

| | Eligibility | Baseline Questionnaire | Randomisation | One-month follow-up | Three-month follow-up | 12-month follow-up |
|---|---|---|---|---|---|---|
| **Consent** | X | X | | X | X | X |
| **Demographic questions** | | X | | | | |
| **PHQ-9**[1] | X | X | | X | X | X |
| **GAD-7** | | X | | X | X | X |
| **De Jong Gierveld Scale (11 items)** | | X | | X | X | X |
| **Lubben Social Network Scale (6 items)** | | X | | X | X | X |
| **SF-12v2** | | X | | X | X | X |

| | | | | | | |
|---|---|---|---|---|---|---|
| **EQ5D-3L** | | X | | X | X | X |
| **Resource use** | | X | | X | X | X |
| **COVID-19 Questions** | | X | | X | X | X |

[1]The PHQ-9 will be administered as part of the screening process and eligibility assessment in order to assess the level of depression symptoms and risk or self-harm/suicide (as per the inclusion and exclusion criteria. Responses are then transferred to the baseline assessment.

#### 6. Outcomes

##### 6.1 Primary outcome

The primary outcome measure is **self-reported depression severity** (as measured by the PHQ-9) and the primary outcome follow-up timepoint is **three months post-randomisation**. This measure is widely used in clinical trials and settings. It provides acceptable internal and external validity and has established specificity/sensitivity in a UK population. [4]. The three-month time point has been chosen as the primary endpoint to test the impact on depression in the medium term and to test whether the intervention mitigates depression during the COVID-19 pandemic period. The BASIL-C19 pilot trial indicated that BA sessions should have been delivered by this post-randomisation point.

The PHQ-9 is the nine-item depression module from the full Patient Health Questionnaire (PHQ). Each of the nine items can be scored from 0 (not at all) to 3 (nearly every day), and a total score is obtained by summing the item scores, to produce a total score ranging from 0-27. A score between 0-4 indicates minimal depression, 5-9 indicates mild depression, 10-14 indicates moderate depression, 15-19 indicates moderately severe depression, and 20-27 indicates severe depression. If one or two item values are missing from the score, then they can be substituted with the average score of the non-missing items (scored pro-rata and total score rounded to nearest integer [5]). Questionnaires with more than two missing values should be disregarded. In the instance where a participant has selected more than one consecutive option for an item, the most severe option will be taken; if two non-consecutive options are selected or more than two options, then the item is treated as missing.

##### 6.2 Secondary outcomes

- **Depression Severity (PHQ-9) –** Self-reported depression severity at one- and 12-months post-randomisation, as measured by the PHQ-9 and detailed in section 6.1.

The following are secondary outcomes at one, three and 12 months:

- **Anxiety (Generalised Anxiety Disorder-7) –** The GAD-7 is a seven-item, self-reported patient questionnaire, which can be used as a screening tool and severity measure for generalised anxiety disorder [6. A total score is calculated by assigning scores of 0, 1, 2 and 3 to the response categories of ‘not at all’, ‘several days’, ‘more than half the days’ and ‘nearly every day’, respectively and adding together the scores for the seven questions. The GAD-7 total score ranges from 0 to 21, with scores of 5, 10 and 15 taken as cut-off points for mild, moderate, and severe anxiety, respectively. If one or two values are missing from the score, then they can be substituted with the average score of the non-missing items (scored pro-rata and total score rounded to nearest integer). Questionnaires with more than two missing values should be disregarded.
- **Loneliness (De Jong Gierveld 11-item Loneliness Scale) –** The De Jong Gierveld Loneliness Scale consists of 11 items, each with three response options: Yes, More or less, and No. Negatively worded questions (items 2, 3, 5, 6, 9, 10) e.g. ‘I miss having a really close friend’ will be coded as Yes = 1, More or less = 1, and No = 0. Positively worded questions (items 1, 4, 7, 8, 11) e.g. ‘There are many people I can trust completely’ will be coded as Yes = 0, More or less = 1, and No = 1. Researchers can use the scale as a one-dimensional measure of loneliness (sum of all items ranging from 0 (not lonely) to 11 (extremely lonely) provided no more than one item is missing) or choose to use two subscales: emotional

loneliness (sum of item 2, 3, 5, 6, 9 and 10; only valid if no missing item data); and social loneliness (sum of items 1, 4, 7, 8 and 11; only valid if no missing item data) [7].

- **Health Related Quality of Life (Short Form-12 version 2)** – The SF-12v2 is a health-related quality of life questionnaire consisting of 12 questions that measure eight health domains to assess physical and mental health. Physical health-related domains include General Health, Physical Functioning, Role Physical and Body Pain. Mental health-related scales include Vitality, Social Functioning, Role Emotional and Mental Health. The physical and mental health component scores both range from 0 to 100, where 0 indicates the lowest level of health and 100 indicates the highest level of health measured by the scale. Scoring will be conducted in accordance with the SF-12v2 scoring manual, via the Optum software[8].
- **Health Related Quality of Life (EuroQol 5 dimension 3 level) –** The EQ5D-3L is a measure of health state (quality of life) comprising five dimensions; mobility, selfcare, usual activities, pain/discomfort and anxiety/depression. Each dimension is scored on a three-point Likert scale: no problems, some problems and unable to do/severe problems. Scores are combined and converted into a summary index value which can be used to facilitate the calculation of quality-adjusted life years (QALYs). This will be analysed as part of the cost-effectiveness analysis.
- **Social Isolation (Lubben Social Network Scale – 6 items) –** This scale is a self-reported measure to gauge social isolation by measuring the number and frequency of social contacts with friends and family members and the perceived social support received from these sources. There are two versions of this scale, with the short 6-item form being utilised here. Each of the six items are scored from 0 – 5, with 0 = none, 1 = one, 2 = two, 3 = three or four, 4 = five to eight and 5 = nine or more. The total score is calculated by finding the sum of all the items (where there are no missing items) and ranges from 0 to 30, with a higher score indicating more social engagement.
- **Circumstances in relation to COVID-19 –** This includes descriptively summarising whether participants are currently social distancing/self-isolating/shielding, number of people living in the household, whether they have felt able to adhere to UK Government guidelines on social distancing/self-isolation, whether they have had COVID-19 and information on the COVID-19 vaccination.

##### 6.3 Other collected variables

The participant’s entry route into the study (direct contact, online consent, or permission to contact will be summarised.

Demographic information will be obtained at baseline and will include a range of sociodemographic variables including age, gender, LTC types/health conditions, socio-economic status, ethnicity, education, cohabitation status, and number of children.

The total number of BA sessions, per participant, and duration in minutes of each session will be recorded. Symptom monitoring at each intervention session will be undertaken using the 7-item depression subscale from the 21-item Depression Anxiety Stress Scale (DASS) [9]. The DASS is widely used/validated in a UK community context and is brief, and simple to score with clear clinical cut off scores (none/mild/moderate/severe). Scores on the DASS will be used to guide decision making by BSWs, guided by supervision provided by clinical members of the BASIL+ study team. Where risk or significant clinical deterioration is noted, the participant would be supported to access more formal healthcare interventions. The DASS will be used during BA sessions as a clinical monitoring tool, and will not be used for analysis purposes.

The study will record details of any Serious Adverse Events (SAEs) experienced by study participants. Adverse Events (AEs) reported by study participants which are not classified as a SAE will be recorded and submitted to the Joint Programme/Trial Steering and Data Monitoring and Ethics Committee in agreement with the Programme/Trial Steering Committee Chair.

The participants' status in the trial will be recorded, and a change in status (i.e. the participant requests to withdraw from the intervention or a participant requests to withdraw fully from the trial) will be recorded, with reasons for the change where provided.

#### 7. Data

##### 7.1 Electronic/non-paper data

Upon receipt of a study information pack in the post, potential participants will be contacted by a member of the practice team (see study protocol for definition). Contact with potential participants will be recorded by the practice team to ensure those patients who decline interest in the study are not contacted again.

For those patients who express an interest in the study during this telephone contact, the practice team will request their verbal 'permission to contact'. The 'permission to contact' will be a verbal agreement for a BASIL+ study researcher to contact them to discuss their suitability for the trial. The 'permission to contact' is not consent to participate. The practice team will provide the BASIL+ study team with the name and contact details of those patients providing their 'permission to contact'; this information will be provided securely (telephone or secure/encrypted email) and recorded.
Alternatively, the Participant Information Sheet (PIS) also offers patients the option to complete an online consent form if they are interested in taking part. The PIS includes a link which directs potential participants to a secure consent form for completion to include essential contact information. Consent form information will be captured via Google Forms, a GDPR-compliant secure software platform – which will be password protected with access limited to named members of the BASIL+ study team. The University of York has a GDPR compliant contract in place with Google GSuite/Cloud to ensure data is handled in accordance with data protection legislation. Completed online consent forms will be received by the central study team. These will be downloaded and then transferred to the relevant site study team by secure/encrypted email, with confirmation of receipt required.
Where an interested patient contacts the BASIL+ study team directly, verbal consent to undertake the eligibility assessment will be obtained and recorded by the study researcher. The BASIL+ study team will liaise with the practice team (via telephone or secure encrypted email) to minimise the risk of these patients being contacted again by the practice team.

Participant personal details (names, addresses, postcodes and other participant contact details) and contact tracking information will be stored electronically using a site-specific 'BASIL+ spreadsheet'. This will be a site-specific Google Sheet or Microsoft Excel Spreadsheet (where access to Google Sheets is not permitted or feasible). The majority of sites have confirmed their ability to access Google Sheets and so the use of Microsoft Excel Spreadsheets will be limited. The Google Sheet or Microsoft Excel Spreadsheet will be held on secure password protected servers at the University of York and at the relevant site organisation (including NHS computers and in secure settings). Access to participant personal details will be restricted to only those members of staff working on the BASIL+ study (e.g. study researchers, site Principal Investigators, BSWs, Trial Administration team). Site teams will only be able to access information for their site participants. Access to site information will be limited to key members of the central study team at the University of York (e.g. study researchers, Chief Investigators, Trial Administration team).

Initial participant details will be added (by site research teams) to an 'Initial Contact and Screening' Google Sheet. A Screening and Eligibility log will be recorded using Google Sheets, or a Microsoft Excel Spreadsheet if a site does not allow use of Google Sheets. This log will include a record of a participant's entry route into the study (permission to contact/online consent/direct contact).

Sites using a Microsoft Excel Spreadsheet will be required to regularly save copies of the spreadsheet with each copy being date stamped. Site study teams will be required to transfer the Microsoft Excel Spreadsheet on an ad hoc basis to the central BASIL+ study team by secure/encrypted mail (with confirmation of receipt).

Data are collected directly from participants using the unique trial identification number (participant ID) allocated at the point of screening. Randomisation via participant ID will involve researcher completion and submission of a randomisation google form (or in liaison with the central study team where site completion of the google form is not feasible).

Participants are telephoned for data collection at baseline, one, three- and 12-months post-randomisation. Data are collected directly into a Qualtrics database by the researcher conducting the phone call. The researcher is blinded to treatment group allocation for the one, three and 12-month follow-up. Qualtrics databases have been created and are managed for this trial by members of the YTU data management team. Qualtrics is a GDPR-compliant survey tool for which the University of York has a site license. Participants opting to complete follow-up outcome measures online will be provided (via email – as provided by the participant) with a secure link to Qualtrics. A new secure link is available upon request if the first link had expired. Data entry via Qualtrics will be via participant ID only and will require double entry of participant ID before data submission.

An ‘Allocation and Intervention’ record will be kept using Google Sheets. Due to blinding, access is restricted to the Trial Manager, the Trial Administration Team and those researchers overseeing intervention delivery. Any researcher that may be involved in conducting follow-up data collection is denied access to this Google Sheet and is blinded to group allocation.

An Intervention Log will be recorded using Google Sheets. The session number and duration (in minutes) will be recorded here, alongside duration of administration time, clinical supervision time and any relevant clinical notes (participant depression scores and intervention notes). If sites do not allow the use of Google Sheets, this data will be recorded using a Microsoft Excel Spreadsheet. Access to these spreadsheets is restricted, with access permitted to only the individual BASIL Support Worker (BSW), the Clinical Supervisors, the BASIL administration team and the Trial Manager. Each individual BSW will be provided with a pre-populated Google Sheet containing participant information (to facilitate participant contact) for their allocated intervention participants only. Each participant will be displayed on separate tabs (or sheets). Where NHS sites have their own systems to record research participant activity, these will be used by site BSWs in place of the aforementioned Google Sheets/Microsoft Excel Spreadsheet. In cases where BSWs use their own systems, the BASIL administration team and the Trial Manager are provided with the relevant information pertaining intervention delivery and the facilitation of clinical supervision. These NHS systems would be research only modules that only BASIL+ site research teams would have access to, no clinical secondary care records would be created for BASIL+ trial participants.

Adverse Events (AEs) and Serious Adverse Events (SAEs) are recorded through Google Forms and Google Sheets. Site researchers, BSWs and Clinical Supervisors complete the Google Form and the data is downloaded to a Google Sheet. This Google Sheet has restricted access and can only be accessed by the Trial Manager, Trial Statistician, Trial Administration team and Clinical Supervisors.

Participant withdrawals are recorded through Google Forms and Google sheets. Site researchers and Clinical Supervisors complete the Google Form and the data is downloaded to a Google Sheet. Access to this Google Sheet is restricted to the Trial Manager, Trial Statistician, Trial Administration team and Clinical Supervisors only.

A log will be kept of researchers who become unblinded during the course of the trial, for example, if a participant discloses information during a telephone follow-up. This log will be recorded using Google Sheets or Microsoft Excel. In the majority of cases, an unblinded researcher would not conduct any subsequent follow-up data collection. In rare cases, an unblinded researcher may conduct subsequent follow-up, if there is a clinical reason to supersede the need for blinding.

At the point of analysis these Google Sheets will be downloaded and saved onto drives at YTU.

#### 8. Sample Size

In an older population with lower severity [subthreshold] depression, a difference of 1.3 PHQ points has been shown to be clinically and cost-effective [10] with a standard deviation of 4, equating to a standardised effect size of 0.325. An effect size of 0.3 is assumed for this calculation, which is also in line with pooled standardised mean differences found in a meta-analysis of previous trials of collaborative care [11]. Assuming 90% power, 5% alpha, 0.3 effect size and 20% attrition (from mortality and loss to follow-up) the trial would need to randomise 590 participants.

However, this calculation is based on a number of conservative assumptions. In this primary analysis, baseline PHQ-9 score shall be included as a covariate, which will result in gains in statistical power. By accounting for the correlation between baseline and outcome, PHQ-9 score leads to a reduction in the required sample size. In the external BASIL-C19 pilot trial, a correlation of 0.58 was observed between the PHQ-9 score at baseline and three months post-randomisation amongst participants who scored ≥5 at baseline, and follow-up at three months was 90%. Assuming a correlation of at least 0.5, 90% power, 5% alpha, 0.3 effect size and 10% attrition, 392 participants would need to be recruited and randomised into the trial. Calculations conducted in STATA v15.

#### 9. Randomisation and Blinding

Following baseline questionnaire completion, eligible and consenting participants will be randomised 1:1 to either receive the intervention (BA) or usual care with signposting information. Randomisation will take place via a secure online randomisation service provided by the York Trials Unit (YTU). A YTU statistician not involved with participant recruitment will generate the allocation schedule, using block randomisation stratified by site with randomly varying block sizes. Participants will be informed of their group allocation by telephone (and confirmed by letter).

Researchers completing follow-up questionnaires (outcome measures) with participants over the telephone will be blind to group allocation. Due to the nature of the intervention, it is not possible to keep participants or BSWs delivering the intervention blind to allocation.

Note. The final number of participants randomised into BASIL+ was 435.

#### 10. Final analysis

##### 10.1 Analysis software

All analyses will be conducted in STATA v17 (StataCorp, 4905 Lakeway Drive, College Station, Texas 77845 USA), or a later version (to be confirmed in final report).

##### 10.2 Analysis principles and populations

Analyses will follow the principles of intention-to-treat with participant’s outcomes analysed according to their original, randomised group, where data are available, irrespective of deviations based on non-compliance. Statistical tests will be two-sided at the 5% significance level and 95% confidence intervals (CIs) shall be used. The statistician will not be blinded to treatment allocation.

##### 10.3 Analysis timelines

There will be two analyses. The first will be conducted following completion of data collection at the three-month (primary) time point. Results of data collected up to and including three months will be published as soon as possible after they are available to avoid delays in the dissemination of the primary outcome findings in view of the prioritisation of research in response to the COVID-19 pandemic and risk of future lockdowns and prolonged isolation for the target population.

At the end of the planned follow up of the trial, the 12-month outcomes of the PHQ-9 (and the secondary outcomes of GAD-7, De Jong Gierveld Scale, Lubben Social Network Scale, and the physical and mental health component scores of the SF-12v2) will then be analysed.

##### 10.4 Screening, eligibility, recruitment and follow-up data

The trial will be reported according to the CONSORT (Consolidated Standards of Reporting Trials Statement) guidelines for a parallel group RCT. The flow of participants through each stage of the trial, including reasons for non-eligibility, will be presented in a CONSORT flow diagram.

The number of individuals screened, eligible and randomised will be presented. Reasons for non-participation will be provided where available. Participants' entry route into the study (permission to contact; direct contact; online consent) will be reported and presented in the CONSORT diagram.

Recruitment graphs presenting the overall recruitment by month, and the actual vs target recruitment will be produced.

The number of participants withdrawing from the intervention and/or the trial will be summarised by trial arm, including timing of withdrawal. Reasons for withdrawal will be provided where available.

The number and proportion of randomised participants followed up at each time point will be reported by treatment group. The mode of data collection (e.g., telephone, online) will be summarised.

##### 10.5 Baseline data

All participant baseline data will be summarised descriptively by trial arm both as randomised and as analysed in the primary analysis. Continuous variables will be reported as means and standard deviations and categorical variables will be reported using counts and percentages. No formal statistical significance testing will be done to test baseline imbalances between the treatment arms, but any noteworthy difference will be descriptively reported.

##### 10.6 Primary analysis

The primary outcome (depression severity as measured by the PHQ-9 at three-months) will be analysed using a linear mixed model, incorporating data from the one and three-month time points. The model will include time, trial arm, an arm-by-time interaction, and baseline PHQ-9 score as fixed effects, with participant nested within site as random effects. The different covariance structures for repeated measurements that are available as part of the analysis software will be applied to the model. The most appropriate pattern will be used for the final model based on diagnostics including Akaike's information criterion (smaller values are preferred). Participants will only be included in the model if they have full data for the baseline covariates and a valid PHQ-9 total score for at least one post-randomisation time point. Missing data are considered missing at random. Model assumptions will be checked as follows: the normality of the standardised residuals will be checked using a QQ plot, and homoscedasticity will be assessed by means of a scatter plot of the standardised residuals against fitted values. If the model assumptions are in doubt, transformations of the outcome data will be considered in sensitivity analyses. A log transformation will be tried in the first instance, and then others as suggested by the Stata 'ladder' command as appropriate. The model will provide a treatment effect at one and three-months, which will be reported as mean differences, 95% confidence intervals and p-values. The primary time-point of interest is pre-specified at three months.

Analysis of the primary outcome will be checked by a second statistician before release of results, and recorded in the YTU *F16: Primary Analysis Sign Off* Form.

##### 10.7 Sensitivity analyses

*Intervention Compliance*

Intervention adherence will be recorded and reported, including the total number of BA sessions completed per participant and the average duration of sessions, which will be summarised descriptively.

A CACE analysis for the primary outcome will be conducted to obtain unbiased estimates of the intervention effectiveness in the presence of full compliance. We will define compliance with the intervention for those allocated to the intervention in two ways: as a continuous measure of the number of BA sessions attended; and as a dichotomous measure to indicate that at least 5 sessions were attended. A two-stage least squares instrumental variable approach will be used (with treatment assignment as the instrumental variable) using the *ivreg* command in Stata, with the *2sls* option [12]. This analysis will use a linear regression model for the PHQ-9 score at 3 months, adjusting for baseline score, and with robust standard errors to account for clustering within site.

##### 10.8 Subgroup analyses

A subgroup analysis will consider whether there is evidence of a differential intervention effect depending on baseline level of depressive symptoms (PHQ-9 score 5-9, or 10 and above). This will be achieved by repeating the primary analysis but including an indicator variable for whether the participant scored 5-9 or 10 or above at baseline as a covariate (rather than the continuous score) plus an interaction term between treatment allocation and baseline PHQ9 threshold.

##### 10.9 Analysis of secondary outcomes

The secondary outcomes of GAD-7, De Jong Gierveld Loneliness Scale (total and two subscales separately), Lubben Social Network Scale, and SF-12 (mental and physical health component scores separately) will be analysed in a similar way to the primary outcome, swapping baseline PHQ-9 score with baseline value of the outcome as a covariate.

Participants' circumstances in relation to COVID-19 at one- and three-months post-randomisation will be summarised descriptively by group using counts and percentages.

##### 10.10 Adverse Events

Serious and non-serious adverse events will be summarised by trial arm and overall, including details of the event, action taken, time to onset, length of event, outcome, relationship to study treatment, and expectedness.

##### 10.11 12-month outcomes

At the end of the trial, the 12-month outcomes of PHQ-9, GAD-7, De Jong Gierveld Loneliness Scale (total and two subscales separately), Lubben Social Network Scale, and SF-12 (mental and physical health component scores separately) will be analysed via a linear mixed model, incorporating data from all post-randomisation timepoints (one, three and 12-months). Only the treatment effects at 12 months and overall will be extracted and reported from these models. Participants' circumstances in relation to COVID-19 at 12-months post-randomisation will be summarised descriptively by group using counts and percentages.

#### 11. Methodological Study Within a Trial (SWAT)

An embedded Study Within a Trial (SWAT) will be undertaken to evaluate the effectiveness of a one-page infographic, to go alongside the Participant Information Sheet (PIS), on participant recruitment and response rates.

This will be tested in a cluster RCT embedded within the BASIL+ trial. Study sites will be randomised 1:1, using simple randomisation, to either the SWAT intervention group (standard study information plus the one-page infographic) or the SWAT control group (standard study information only). Generation of the SWAT allocation sequence will be undertaken independently by a statistician at YTU not involved with the BASIL+ recruitment process. Allocation to either SWAT group is entirely separate and independent from subsequent individual participant randomisation into the BASIL+ main trial.

Since this is a SWAT, formal power calculation has not been conducted. The sample size for the SWAT will be determined by the number of sites opened to recruitment and the number of potential participants approached.

The primary outcome of this embedded SWAT will be the participant recruitment rate into the BASIL+ trial. Secondary outcomes will include:

i. Participant response rate in terms of expressions of interest (permission to contacts received, direct participant contacts, or participant completion of the online consent form);
ii. Retention in the main trial as measured by response to the three-month post-randomisation participant questionnaire.

The proportion of participants randomised into the main BASIL+ trial will be compared between the SWAT groups using a mixed-effect logistic regression model, adjusting for trial site as a random effect. The secondary outcomes will be similarly analysed; the model for response rate at the three-month follow-up will also include a covariate for main trial allocation (intervention/usual care/not randomised).

#### 12. SAP amendment log

All changes that are made to the Statistical Analysis Plan following initial sign-off will be detailed in the box below.

| Amendment/addition to SAP and reason for change | New version number, name and date |
|---|---|

#### 13. Signatures of approval

Sign-off of the final approved version of the Statistical Analysis Plan by the principle investigator and trial statistician(s) (can also include Trial Manager/Co-ordinator)

| Name | Trial Role | Signature | Date |
|---|---|---|---|
| Prof Simon Gilbody | Co-chief investigator | 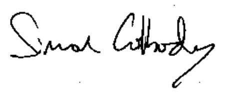 | Twenty-sixth of September two thousand and twenty-two |
| Prof David Ekers | Co-chief investigator | 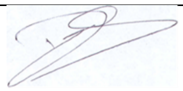 | Twenty-sixth of September two thousand and twenty-two |

| | | | |
|---|---|---|---|
| Dr Liz Littlewood | Trial/Programme Manager | [illegible] | Twenty-sixth of September two thousand and twenty-two |
| Kalpita Baird | Statistician | Kalpita Baird | Twenty-sixth of September two thousand and twenty-two |
| Caroline Fairhurst | Senior Statistician | C.M.Fairhurst | Twenty-sixth of September two thousand and twenty-two |
